# Head and Neck Cancer Primary Tumor Auto Segmentation using Model Ensembling of Deep Learning in PET-CT Images

**DOI:** 10.1101/2021.10.14.21264953

**Authors:** Mohamed A. Naser, Kareem A. Wahid, Lisanne V. van Dijk, Renjie He, Moamen Abobakr Ab-delaal, Cem Dede, Abdallah S.R. Mohamed, Clifton D. Fuller

## Abstract

Auto-segmentation of primary tumors in oropharyngeal cancer using PET/CT images is an unmet need that has the potential to improve radiation oncology workflows. In this study, we develop a series of deep learning models based on a 3D Residual Unet (ResUnet) architecture that can segment oropharyngeal tumors with high performance as demonstrated through internal and external validation of large-scale datasets (training size = 224 patients, testing size = 101 patients) as part of the 2021 HECKTOR Challenge. Specifically, we leverage ResUNet models with either 256 or 512 bottleneck layer channels that are able to demonstrate internal validation (10-fold cross-validation) mean Dice similarity coefficient (DSC) up to 0.771 and median 95% Hausdorff distance (95% HD) as low as 2.919 mm. We employ label fusion ensemble approaches, including Simultaneous Truth and Performance Level Estimation (STAPLE) and a voxel-level threshold approach based on majority voting (AVERAGE), to generate consensus segmentations on the test data by combining the segmentations produced through different trained cross-validation models. We demonstrate that our best performing ensembling approach (256 channels AVERAGE) achieves a mean DSC of 0.770 and median 95% HD of 3.143 mm through independent external validation on the test set. Concordance of internal and external validation results suggests our models are robust and can generalize well to unseen PET/CT data. We advocate that ResUNet models coupled to label fusion ensembling approaches are promising candidates for PET/CT oropharyngeal primary tumors auto-segmentation, with future investigations targeting the ideal combination of channel combinations and label fusion strategies to maximize segmentation performance.

## 1 Introduction

Oropharyngeal cancer (OPC) is a type of head and neck squamous cell carcinoma that affects a large number of individuals across the world [1]. Radiation therapy is an effective component of OPC treatment but is highly dependent on accurate segmentation of gross tumor volumes [2], i.e., visible gross disease that is informed by clinical examination and radiographic findings. Importantly, precise tumor delineation is crucial to ensure adequate radiation therapy dose to target volumes while minimizing dose to surrounding healthy tissues. The combination of computed tomography (CT) with positron emission tomography (PET) allows for sufficient anatomic detail in determining tumor location coupled to underlying physiologic information [3]. However, tumor segmentation in OPC has long been seen as an inefficient and potentially inconsistent process as multiple studies have demonstrated high inter- and intra-observer segmentation variability [4, 5]. Therefore, developing automated tools to reduce the variability in OPC PET-CT tumor segmentation while retaining reasonable performance is imperative for improving the radiation therapy workflow.

The annual Medical Image Computing and Computer Assisted Intervention Society (MICCAI) Head and Neck Tumor Segmentation Challenge (HECKTOR) has provided an avenue to systematically evaluate different OPC primary tumor auto-segmentation methodologies through the release of high-quality, multi-institutional training and testing PET-CT data. We previously participated in the 2020 HECKTOR challenge and achieved reasonable results using deep learning approaches [6]. Subsequently, we improve upon our previous approach through various architectural modifications, ensembling of independent models’ predictions, and additional training data provided by the Challenge that ultimately leads to improved segmentation performance. This workpresents the results of our OPC primary tumor auto-segmentation model based on a ResUnet deep learning model applied to the 2021 HECKTOR Challenge PET-CT training and testing data.

## 2 Methods

We developed a deep learning model (2.3) for auto-segmentation of primary tumors of OPC patients using co-registered ^18^F-FDGPET and CT imaging data (2.1). The ground truth manual segmentation of the tumors and the normalized imaging data (2.2) were used to train the model (2.4). The performance of the trained model for auto-segmentation was validated usinga 10-fold cross-validation approach (2.5).

### 2.1 Imaging Data

The data set used in this study, which was released through AIcrowd [7] for the HECKTOR Challenge at MICCAI 2021 [8], consists of co-registered ^18^F-FDG PET and CT scans for 325 OPC patients (224 patients used for training and 101 patients used for testing, previously partitioned by the HECKTOR Challenge organizers). All imaging data in the training set (224 patients) was paired with ground truth manual segmentations of the OPC primary tumors derived from clinical experts (HECKTOR Challenge organizers). All training and testing data were provided in Neuroimaging Informatics Technology Initiative (NIfTI) format.

### 2.2 Image Processing

All images (i.e., PET, CT, and tumor segmentation masks) were cropped to fixed bounding box volumes, provided with the imaging data (2.1) by [7], of size 144×144×144 mm^3^ in the x, y and z dimensions. To mitigate the variable resolution of the PET and CT images, the cropped images were resampled to a fixed image resolution of 1 mm in the x, y, and z dimensions. We used spline interpolation of order 3 for resampling the PET/CT images and nearest-neighbor interpolation for resampling the segmentation masks. We based our cropping and resampling work on the code provided by the HECKTOR Challenge organizers (https://github.com/voreille/hecktor). The CT intensities were truncated in the range of [-200, 200] Hounsfield Units (HU) to increase soft tissue contrast and then were normalized to a [-1, 1] scale. The intensities of PET images were normalized with z-score normalization ([intensity-mean]/standard deviation). We used the Medical Open Network for AI (MONAI) [9] software transformation packages to rescale and normalize the intensities of the PET/CT images. Image processing steps used in this manuscript are displayed in **Figure 1**.

**Fig. 1.**
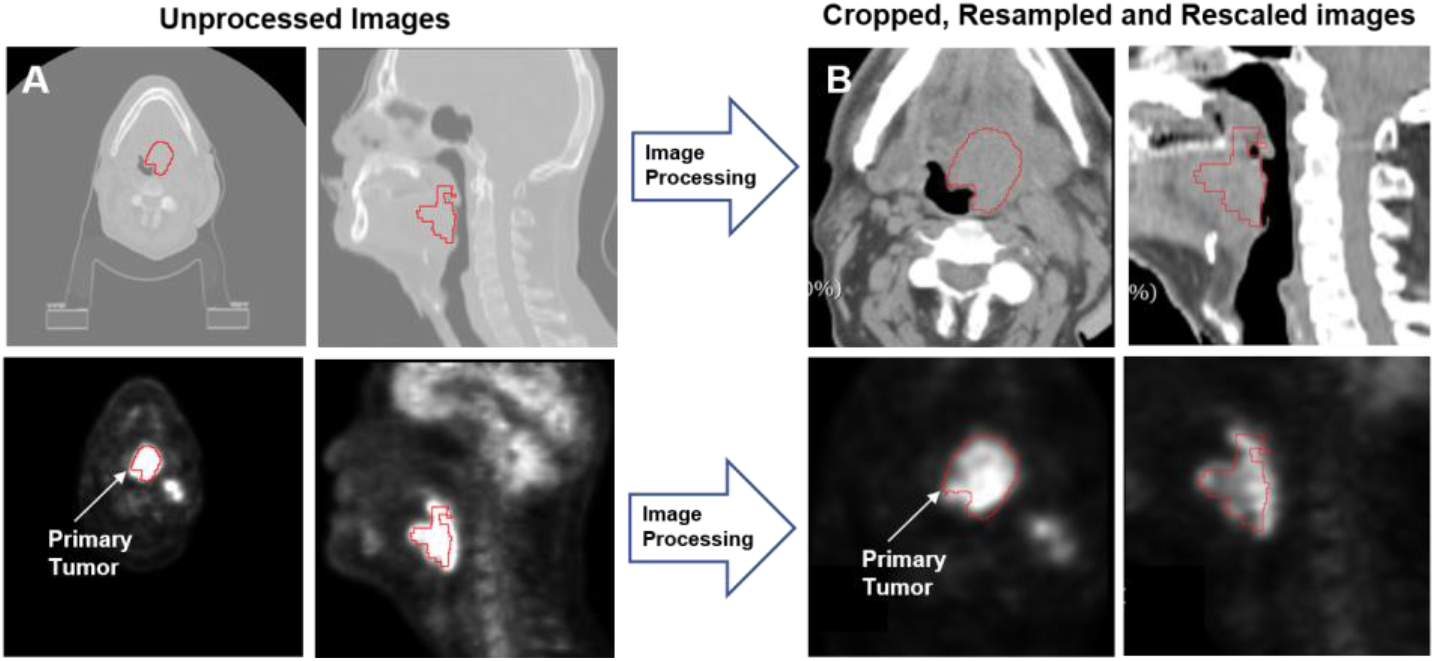
An illustration of the workflow used for image processing. (A) Overlays of the provided ground truth tumor segmentation mask and the original CT (top) and PET (bottom) images. (B) Overlays of the provided ground truth tumor segmentation masks and the processed CT (top) and PET (bottom) images.

### 2.3 Model Architecture

A deep learning convolutional neural network model based on the ResUnet architecture included in the MONAI software package was used for the analysis. As shown in **Figure 2**, the network consists of 4 convolution blocks in the encoding and decoding branches and a bottleneck convolution block between the two branches. All convolution layers use a kernel size of 3 except one convolution layer in the bottleneck, which uses a kernel size of 1. The number of output channels for each convolution layer is given above each layer, as shown in **Figure 2**. Each convolution block in the encoding branch is composed of a two-strided convolution layer and a residual connection that contains a two-strided convolution layer and a one-strided convolution layer. In the bottleneck, the residual connection contains two one-strided convolution layers. In the decoding branch, each block contains a two strided convolution transpose layer, a one strided convolution layer and a residual connection. Batch normalization and Parametric ReLU (PReLU) activation functions were used throughout the architecture. The PET/CT images acted as two channel inputs to the model, and a two-channel output to provide the tumor segmentation mask (i.e., 0 = background, 1 = tumor). The architecture shown in **Figure 2** is for a ResUnet with a maximum number of channels in the bottleneck layer of 512 (512 Model) and the number of channels in the convolution layers of (32, 64, 128, 256, and 512). We also implemented a model using a maximum of 256 channels in the bottleneck layer (256 Model), which has the same structure as the 512 Model, but the number of channels used were (16, 32, 64, 128, and 256).

**Fig. 2.**
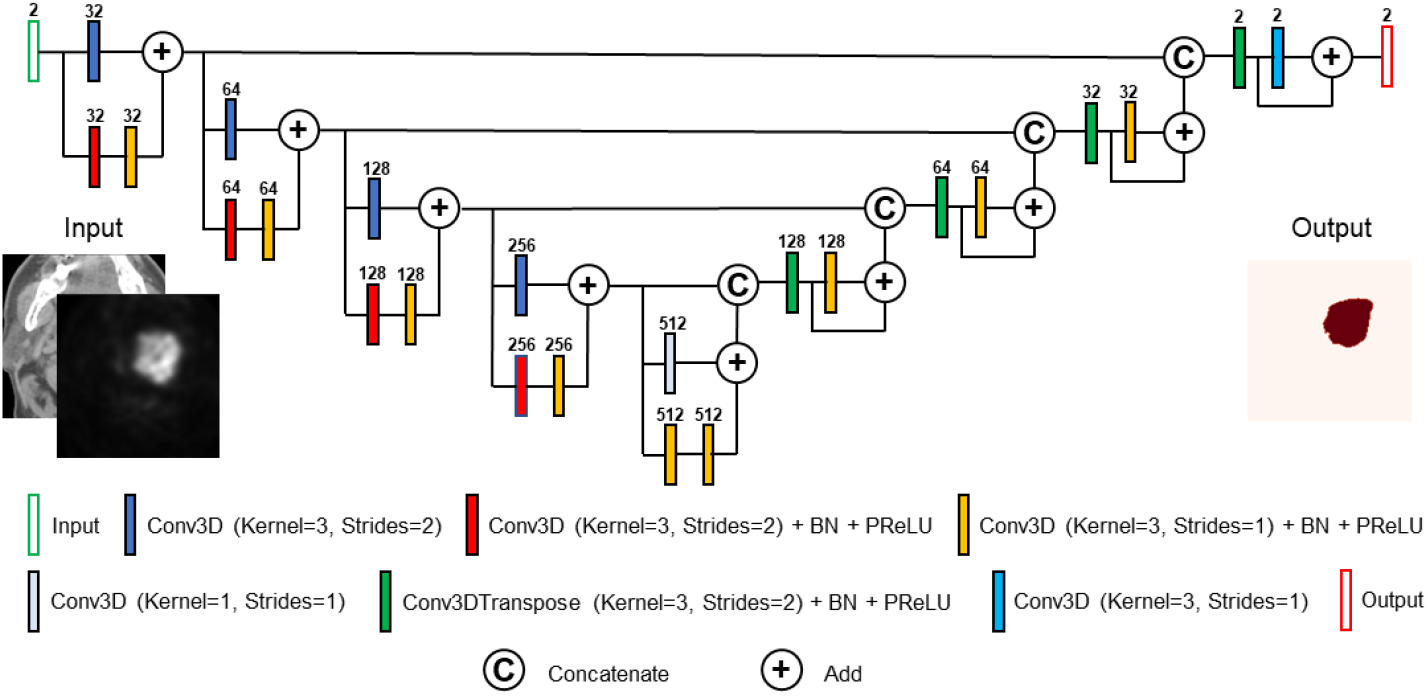
Schematic of the ResUnet architecture used for the segmentation model. The number of channels (32, 64, 128, 256, and 512) is given above each block. The batch normalization and the parametric ReLU layers are annotated by (BN) and PReLU, respectively. The channels given in the figure are for the 512 model, while for the 256 model the channels are (16, 32, 64, 128, and 256).

### 2.4 Model Implementation

We used a 10-fold cross-validation approach where the 224 patients from the training data were divided into 10 non-overlapping sets. Each set (22 patients) was used for model validation while the remaining 202 patients in the remaining sets were used for training, i.e., each set was used once for testing and 9 times for training. The processed PET, CT, and tumor masks (2.2) were randomly cropped to four random fixed-sized regions (patches) of size (96, 96, 96) per patch per patient. The random spatial cropping considered the patch center of mass as foreground (i.e., a tumor positive) or background (i.e., non-tumor - negative) with a 50% probability for both the positive and negative cases as shown in **Figure 3A**. We used a batch size of 2 patients’ images and, therefore, a total of 8 patches of images. The shape of the input tensor provided to the network (2.3) for a batch size of 2, patches per image of 4, a two-channel input (PET/CT), and patch size of (96, 96, 96) is (8, 2, 96, 96, 96). The tumor mask was used as the ground truth target to train the segmentation model. The shape of the target tensor provided was (8, 1, 96, 96, 96). To minimize overfitting, in addition to the random spatial cropping to patch the images and masks, we implemented additional data augmentation to both image and mask patches which includes random horizonal flips of 50%, and random affine transformations with an axial rotation range of 12 degrees and scale range of 10%. We used Adam as the optimizer and Dice loss as the loss function. The model was trained for 700 iterations with a learning rate of 2×10^−4^ for the first 550 iterations and 1×10^−4^ for the remaining 150 iterations. The image processing (2.2), data augmentation, network architecture, and loss function were used from the software packages provided by the MONAI framework [9]; code for these packages can be found at “https://github.com/Project-MONAI/“.

**Fig. 3.**
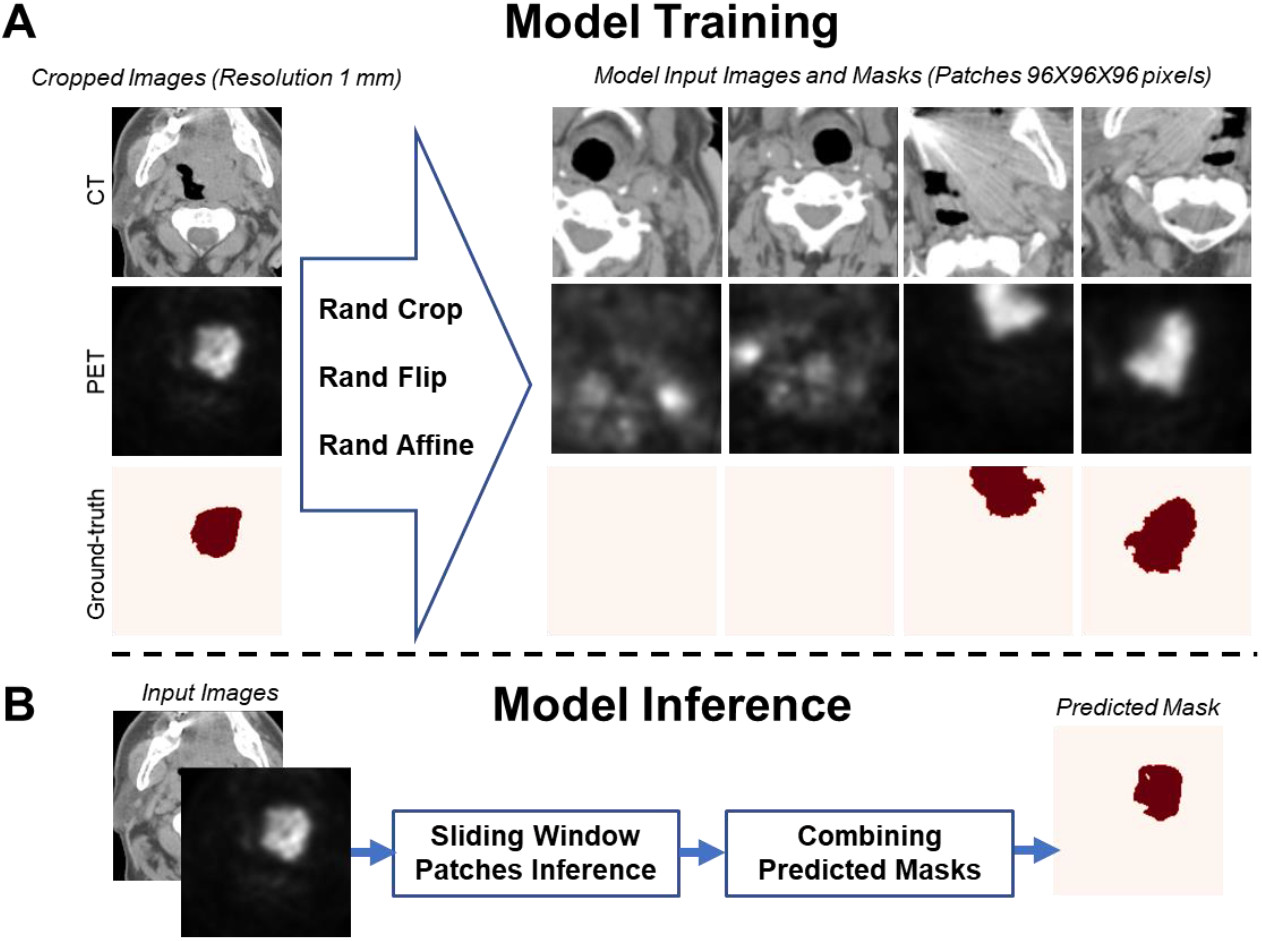
An illustration of the workflow of the training and inference phases of the segmentation model. (A) Data transformation and augmentation is used to produce input data to the model. An example of four patches of CT and PET images and the corresponding ground truth tumor segmentation with at 50% representation of the tumor in these patches used for training the segmentation model (four patches of images per patient - 96 × 96 × 96 voxels each). (B) Segmentation model prediction using sliding window inferences (96×96×96 voxels each) and combining the predicted masks from all patches to provide the final mask.

### 2.5 Model Validation

For each validation fold (i.e., 22 patients), we trained the 256 and 512 ResUnet models (2.3) on the remaining 202 patients. Therefore, we obtained 10 different models for the 256 and 512 networks each from 10-fold cross-validation. We applied an argmax function to the two-channel output of each model to generate the predicted tumor segmentation mask (i.e., 0 = background, 1 = tumor). We evaluated the performance of each separate model on the corresponding validation set using metrics of spatial overlap (Sørensen–Dice similarity coefficient [DSC] [10], recall, and precision) and surface distance (surface DSC [11], 95% Hausdorff distance [95% HD] [12]) between generated and ground truth segmentations. The surface distance metrics were calculated using the surface-distances Python package by DeepMind [11]. A tolerance of 3.0 mm was chosen for calculation of surface DSC based on previous investigations [13, 14] as a reasonable estimate of human inter-observer error.

For the test set (101 patients), we implemented two different model ensembling approaches post-hoc (after training) to estimate the predicted tumor masks. In the first approach, we use used the Simultaneous Truth and Performance Level Estimation (STAPLE) algorithm [15] as a method to fuse labels generated by applying the 10 models produced during the 10 fold cross-validation on the test data set, i.e., generate the consensus predicted masks from the different generated predicted masks (STAPLEapproach). The STAPLE algorithm was derived from publicly available Python code(https://github.com/fepegar/staple). In the second approach, we took a simple threshold of agreement based on all cross-validation fold models at the voxel-level (AVERAGE approach). The total number of cross-validation models used in thresholding could be modulated as a paremeter for this approach. For our purposes, we selected a threshold of 5 cross-validation folds as a proxy for majority voting, i.e. at least 5 cross validation models must consider a voxel to be a tumor (label = 1) for that final voxel label to be considered as a tumor (label =1). Majority voting in this context was chosen since it is common in other model ensembling approaches [16].

## 3 Results

The performance of the segmentation models is illustrated in **Fig. 4**, which shows Box-plots of the DSC, recall, precision, surface DSC, and 95% HD distributions obtained using the 10-fold cross-validation approach described in (2.5). The mean ± standard deviation values of the DSC, recall, precision, surface DSC, and 95% HD achieved by the 256 and 512 Models are 0.771 ± 0.039 and 0.768 ± 0.041, 0.807 ± 0.042 and 0.793 ± 0.038, 0.788 ± 0.038 and 0.797 ± 0.038, 0.892 ± 0.042 and 0.890 ± 0.044, and 6.976 ± 2.405 and 6.807 ± 2.357 respectively. The mean and median values of these metrics are summarized in **Table 1**. Notably, one case did not return a segmentation prediction (CHUS028) for either the 256 or 512 models, which led to the spurious prediction of surface distance metrics. Therefore, this case has been excluded in the analysis of surface DSC and 95% HD.

**Table 1.**
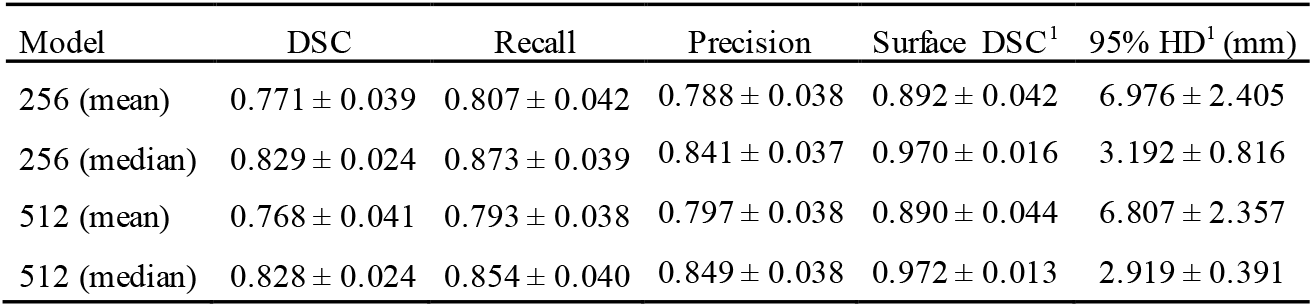
256 and 512 ResUnet model performance metrics. ^1^ One case (CHUS028) in a cross-validation fold contained no segmentation prediction for either model and led to erroneous surface distance calculations; therefore, this case was excluded from the presented surface distance metric results.

| Model | DSC | Recall | Precision | Surface DSC<sup>1</sup> | 95% HD<sup>1</sup> (mm) |
| --- | --- | --- | --- | --- | --- |
| 256 (mean) | $0.771 \pm 0.039$ | $0.807 \pm 0.042$ | $0.788 \pm 0.038$ | $0.892 \pm 0.042$ | $6.976 \pm 2.405$ |
| 256 (median) | $0.829 \pm 0.024$ | $0.873 \pm 0.039$ | $0.841 \pm 0.037$ | $0.970 \pm 0.016$ | $3.192 \pm 0.816$ |
| 512 (mean) | $0.768 \pm 0.041$ | $0.793 \pm 0.038$ | $0.797 \pm 0.038$ | $0.890 \pm 0.044$ | $6.807 \pm 2.357$ |
| 512 (median) | $0.828 \pm 0.024$ | $0.854 \pm 0.040$ | $0.849 \pm 0.038$ | $0.972 \pm 0.013$ | $2.919 \pm 0.391$ |

**Fig. 4.**
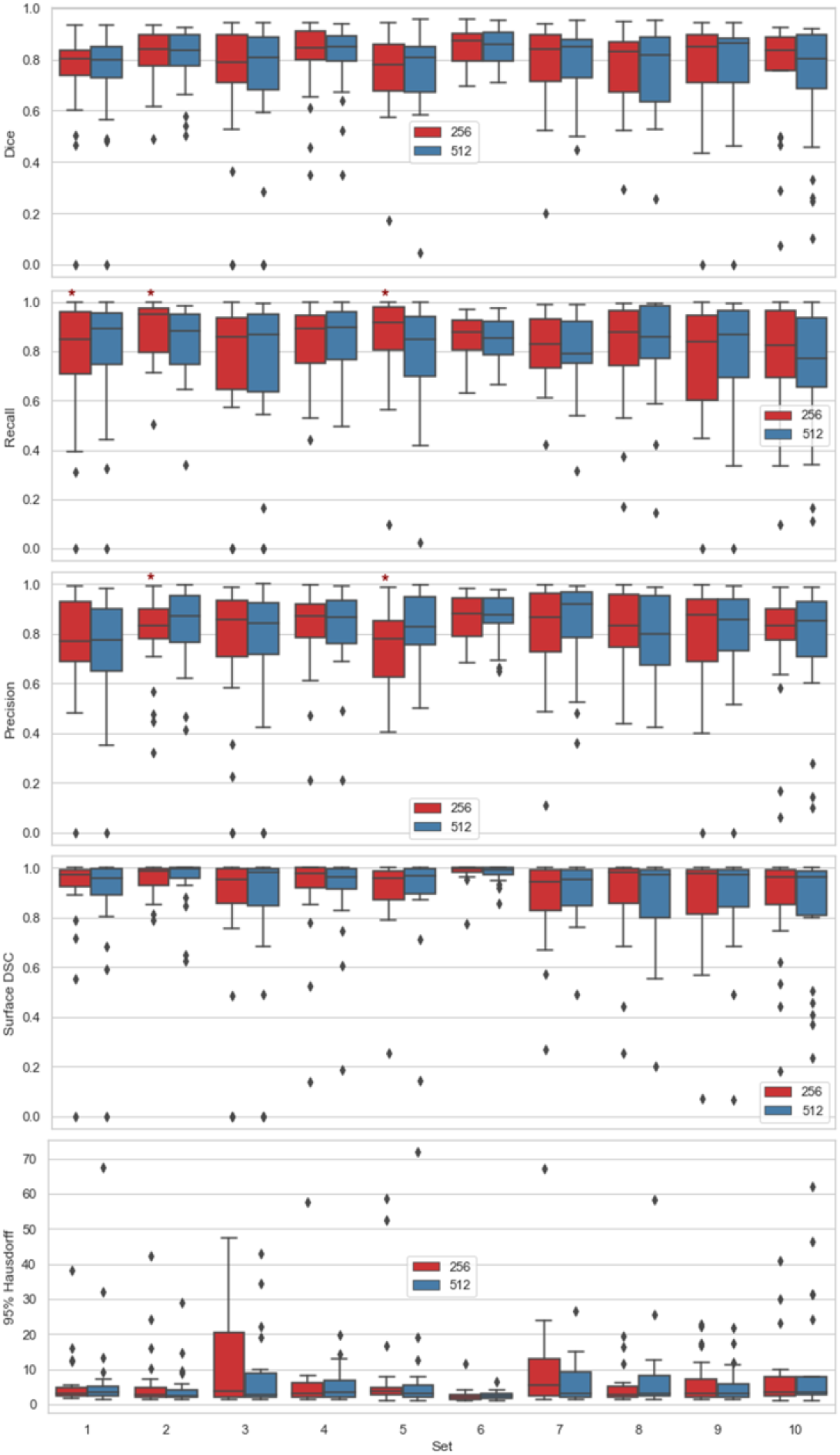
Boxplots of the DSC, recall, precision, surface DSC, and 95% HD distributions for the 10-fold cross-validation data sets (Set 1 to Set 10 – 22 patients each*) used for the 256 and 512 ResUnet models. The lines inside the boxes refer to the median values. The stars refer to significant differences in the results by the two models (p-value < 0.05) using two-sided Wilcoxon signed-rank test. ^1^ One patient in Set 1 (CHUS028) did not return a segmentation prediction for either model and was thus excluded from the analysis of surface distance metrics (surface DSC, 95% HD).

To visually illustrate the internal validation performance of the segmentation model, samples of overlays of CT and PET images with the outlines of tumor masks using ground truth and model segmentations from the validation data sets are shown in **Fig. 5**. The figure shows representative segmentation results for DSC values of 0.54, 0.77, and 0.96 which are below, comparable, and above the segmentation model’s mean DSC of 0.77, respectively.

**Fig. 5.**
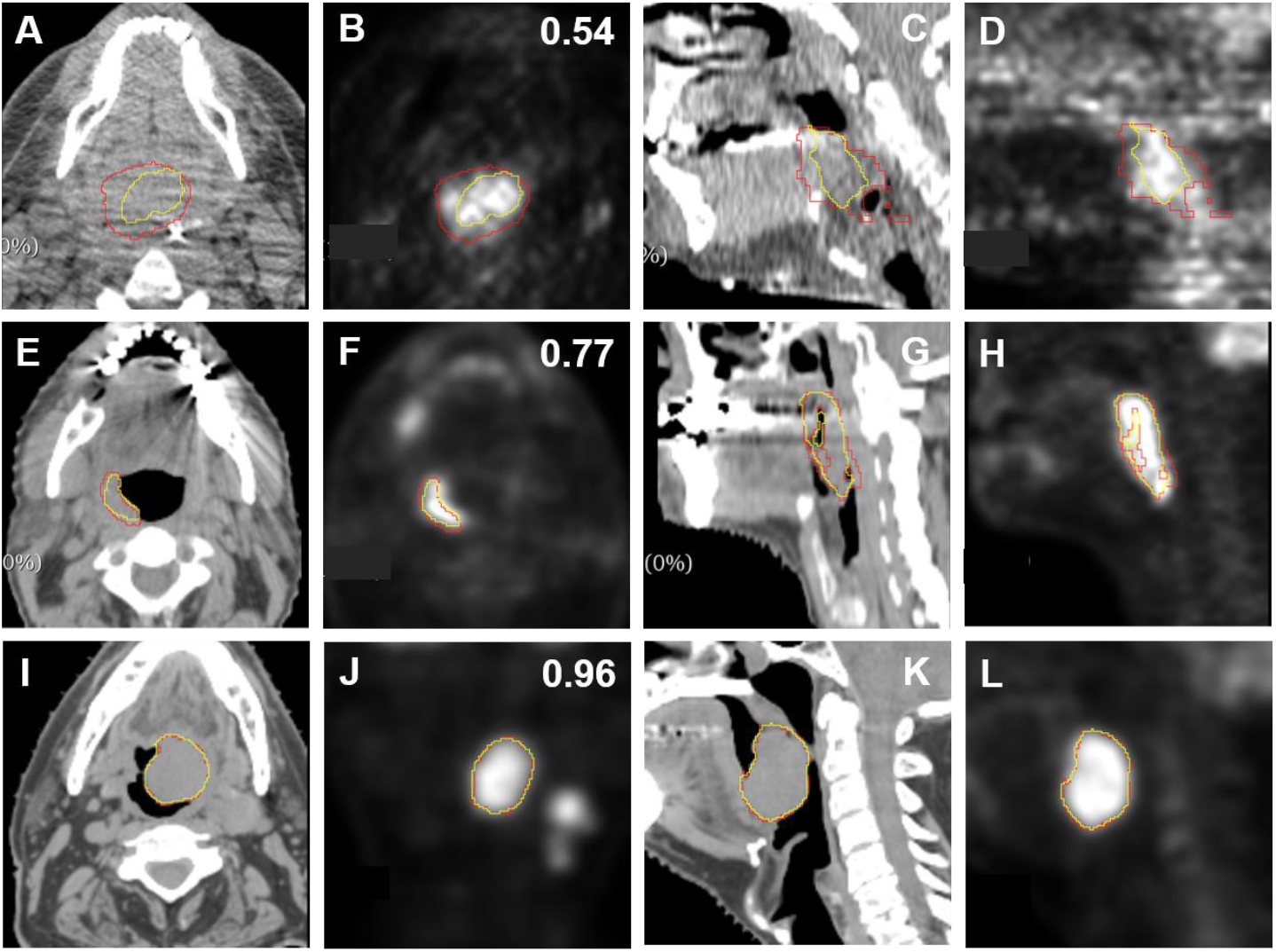
Illustrative examples overlaying the ground truth tumor segmentations (red) and predicted tumor segmentations (yellow) on the CT images (first and third columns) and PET images (sec-ond and forth columns) with different 3D volumetric DSC values (below, equivalent, and above the mean estimated DSC value of 0.77) given at the right top corners of the PET images in the second column.

Finally, our models’ external validation performance (test set) based on ensembling of cross-validation folds previously described are shown in **Table 2**. Mean DSC and median 95% HD for our best model (256 AVERAGE) was 0.770 and 3.143 mm, respectively (standard deviation or confidence intervals not provided by the HECKTOR 2021 submission portal).

**Table 2.**
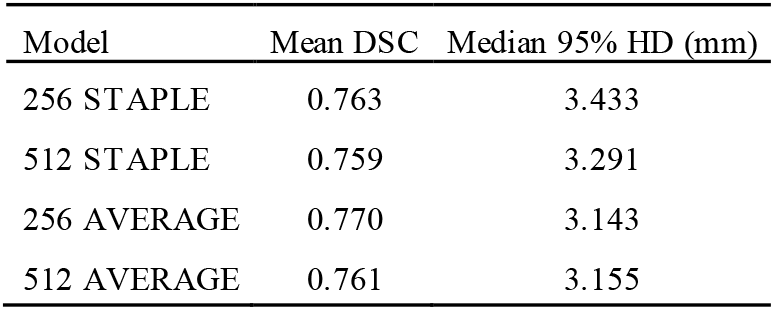
Test set results for ensemble models. Metrics were derived from the HECKTOR 2021 submission portal.

| Model | Mean DSC | Median 95% HD (mm) |
| --- | --- | --- |
| 256 STAPLE | 0.763 | 3.433 |
| 512 STAPLE | 0.759 | 3.291 |
| 256 AVERAGE | 0.770 | 3.143 |
| 512 AVERAGE | 0.761 | 3.155 |

## 4 Discussion

In this study, we have trained and evaluated OPC primary tumor segmentation models based on the 3D ResUnet deep learning architecture applied to large-scale and multi-institutional PET-CT data from the 2021 HECKTOR Challenge. Moreover, we investigate a variety of architectural modifications (512 vs. 256 channels in bottleneck layer) and ensembling techniques (STAPLE vs. AVERAGE) for test set predictions. Our approach yields high and consistent segmentation performance in internal validation (cross-validation) and external validation (independent test set), thereby providing further empirical evidence for the feasibility of deep learning-based primary tumor segmentation for fully-automated OPC radiotherapy workflows.

Through internal validation procedures on the training set (10-fold cross-validation), we attain mean DSC, recall, and precision values of 0.771, 0.807, and 0.788 for the 256 model and 0.768, 0.793, and 0.797, for the 512 model, respectively. While the 512 model offers a greater number of channels that could provide greater contextual information, maximum DSC performance is achieved with the 256 model. This may indicate the 256 model led to less over-fitting on the training data evaluation procedure. Interestingly, there was a tradeoff between recall and precision for the two models tested, with the 256 model offering higher recall at the cost of precision compared to the 512 model. Regardless, both these internal validation results improve upon our 3D models implemented in the 2020 HECKTOR Challenge, which only achieved a mean DSC of 0.69 [6]. These improved results potentially highlight the utility and importance of residual connections in a Unet architecture for this task. Moreover, image processing approaches that improve target class balance (i.e., tumor vs non-tumor) in the provided images significantly improve model sensitivity. Finally, we have further investigated the performance of our models using surface distance metrics, as these metrics have been suggested to be more closely linked to clinically meaningful endpoints [17, 18]. We observe minimal differences between the two models for the surface DSC, with both models showing strong performance. However, the 512 model has a slightly lower 95% HD, which may be favorable when more precise tumor boundary definitions are desired.

When models were evaluated on the test data (external validation), we demonstrate high performance consistent with the internal validation results. Generally, the AVERAGE method outperformed the STAPLE method in terms of both DSC and HD. Typica **l**y, the AVERAGE method led to more conservative estimates than the STAPLE method, which could indicate ground truth segmentations in the test set tended to be more conservative when considering tumor boundaries. Interestingly, while a tradeoff between DSC and HD exists based on the channel number for the STAPLE method (256 = better DSC, 512 = better HD), this tradeoff is not present with the AVERAGE method, as the 256 model has better DSC and HD compared to the 512 model. Compared to our entry for the 2020 HECKTOR Challenge [6], our mean DSC test results were improved by a sizable degree from the original DSC of 0.637 (0.133 increase for our best model). Moreover, we also improve upon the performance of the winning submission in the 2020 HECKTOR Challenge [19], which achieved a DSC of 0.759 (0.011 increase for our best model). Our positive results may in part be due to the inclusion of ensembling coupled to our improved network modifications (as indicated in the internal validation). The utility of ensembling for PET-CT OPC tumor segmentation has been previously noted since the winning entry in the 2020 challenge used an ensembling approach based on leave-one-center-out cross-validation models to yield the best performing DSC results [19]. Therefore, our results further incentivize the ensembling of model predictions for OPC tumor segmentation data.

In recent years, there has been increasing evidence suggesting the utility of applying deep learning for fully-automated OPC tumor auto-segmentation in various imaging modalities [14, 20–22]. PET-CT has recently shown excellent performance when used as inputs to deep learning models, partly due to the large and highly curated datasets provided by the HECKTOR Challenge [8]. While direct comparison of performance metrics between segmentation studies is often ill-advised, the HECKTOR Challenge offers a systematic method for directly compare segmentation methods with each other. Moreover, since it has been suggested that the mean interobserver DSC for head and neck tumors in human experts is approximately 0.69 [23], our results indicate the potential for further testing to develop auto-segmentation workflows. However, it should be noted that before any definitive statements could be said about the clinical value of an auto-segmentation tool, the dosimetric impact and clinical acceptability of auto-segmeneted structures should be thoroughly evaluated through further studies [18].

One limitation of our study is the reliance of our loss function purely on the DSC as an optimization metric. We have chosen the DSC loss since it has provided excellent results in previous investigations and due to its general ubiquity. However, other loss functions such, as cross entropy [6] and focal loss [19], can be combined with the DSC loss for model optimization which may require further invetigation. Moreover, additional measures of spatial similarity, such as surface DSC and 95% HD, are relevant in auto-segmentation for radiotherapy applications [18], and therefore may be attractive candidates for use in model loss optimization [24]. The importance of additional measures of spatial similarity seems to have been noted by the HECKTOR Challenge organizers, as the 95% HD has now become a metric used in the leaderboard to rank contestant performance. An additional limitation of our study is we have only tested a few label fusion approaches as ensembling techniques for our models. For example, we have selected STAPLE as a label fusion method because of its general ubiquity and widely available implementations. However, STAPLE has been criticized in the past [25]; therefore, additional label fusion approaches may be necessary to test in this framework [26]. Moreover, for the AVERAGE ensembling method, the specific threshold in the number of cross-validation models used to determine final label fusion can be seen as an additional parameter to tune. While we have chosen a 5 model threshold as a proxy for majority voting, alternative thresholding strategies can lead to more conservative or liberal estimates of tumor segmentation.

## 5 Conclusion

This study presented the development and validation of deep learning models using a 3D ResUnet architecture to segment OPC primary tumors in an end-to-end automated workflow based on PET-CT images. Using a combination of pre-processing steps, architectural design decisions, and model ensembling approaches, we achieve promising internal and external validation segmentation performance, with external validation mean DSC and median 95% HD of 0.770 and 3.143 mm, respectively, for our best model. Our method notably improves upon our previous iteration of our model submitted in the 2020 HECKTOR Challenge. Future studies should seek to further optimize these methods for improved OPC tumor segmentation performance in forthcoming iterations of the HECKTOR Challenge.

## Data Availability

Data are available from the MICCAI HECKTOR 2021 Challenge Organizers.

## Acknowledgements

M.A.N. is supported by a National Institutes of Health (NIH) Grant (R01 DE028290-01). K.A.W. is supported by a training fellowship from The University of Texas Health Science Center at Houston Center for Clinical and Translational Sciences TL1 Program (TL1TR003169), the American Legion Auxiliary Fellowship in Cancer Research, and a NIDCR F31 fellowship (1 F31 DE031502-01). C.D.F. received funding from the National Institute for Dental and Craniofacial Research Award (1R01DE025248-01/R56DE025248) and Academic-Industrial Partnership Award (R01 DE028290), the National Science Foundation (NSF), Division of Mathematical Sciences, Joint NIH/NSF Initiative on Quantitative Approaches to Biomedical Big Data (QuBBD) Grant (NSF 1557679), the NIH Big Data to Knowledge (BD2K) Program of the National Cancer Institute (NCI) Early Stage Development of Technologies in Biomedical Computing, Informatics, and Big Data Science Award (1R01CA214825), the NCI Early Phase Clinical Trials in Imaging and Image-Guided Interventions Program (1R01CA218148), the NIH/NCI Cancer Center Support Grant (CCSG) Pilot Research Program Award from the UT MD Anderson CCSG Radiation Oncology and Cancer Imaging Program (P30CA016672), the NIH/NCI Head and Neck Specialized Programs of Research Excellence (SPORE) Developmental Research Program Award (P50 CA097007) and the National Institute of Biomedical Imaging and Bioengineering (NIBIB) Research Education Program (R25EB025787). He has received direct industry grant support, speaking honoraria and travel funding from Elekta AB.

## Notes

### Competing Interest Statement

The authors have declared no competing interest.

### Author Declarations

All data was used with approval from the HECKTOR Challenge organizers. Institutional Review Boards (IRB) of all participating institutions permitted the use of images and clinical data, either fully anonymized or coded, from all cases for research purposes, only. Retrospective analyses were performed following the relevant guidelines and regulations as approved by the respective institutional ethical committees with protocol numbers: MM-JGH-CR15-50 (HGJ, CHUS, HMR, CHUM) and CER-VD 2018-01513 (CHUV).

